# Lost Opportunity in Tobacco Cessation Care: Impact of Underbilling in a Large Health System

**DOI:** 10.1101/2024.03.29.24304678

**Authors:** Derek J Baughman, Marcus Rauhut, Edward Anselm

## Abstract

**Introduction:** Tobacco cessation remains a critical challenge in healthcare, with evidence-based interventions often under-utilized due to misaligned economic incentives and inadequate training. This study aims to quantify the economic impact of missed billing opportunities for tobacco cessation in a healthcare system, thereby assessing potential revenue loss and evaluating the effectiveness of systems-based approaches in enhancing tobacco cessation efforts.

**Methods:** A retrospective cohort study utilized aggregated de-identified patient health data from an 8-hospital regional health system across Pennsylvania and Maryland, from 1/1/21 to 12/31/23. The analysis focused on primary care encounters eligible for tobacco cessation counseling (CPT codes 99406 or 99407), with potential revenue calculated based on the Medicare reimbursement rate.

**Results:** Over three years, and 507,656 office visits, only 1,557 (0.3%) of encounters with persons using tobacco were billed for cessation services. The estimated total potential revenue gained if each person who was identified as using tobacco was billed consistently for tobacco cessation counseling was $5,947,018.13, and $1,982,339.38 annually.

**Conclusions:** The study reveals a significant gap between the potential and actual billing for tobacco cessation services, highlighting not only the financial implications of missed opportunities but the validation of health system’s public health impact. Underbilling contributes to considerable annual revenue loss and undermines primary prevention efforts against tobacco-related diseases. Our findings illuminate the need for enhanced billing practices and systemic changes, including policy improvements that influence proper billing to promote public health benefits through improved tobacco cessation interventions.

## Introduction

Despite the progress in reducing the prevalence of adult smoking in the United States, which is now at 11.5%, there are still over 28.3 million adult persons using cigarettes who could benefit from quitting.^1^ The US prevalence of all forms of tobacco use combined is estimated at 18.7%.^1^ According to the recent Surgeon General’s Report on Smoking Cessation (2020),^2^ 77% of people who smoke express interest in quitting, with 55.4% of persons who smoke making a quit attempt in the previous year. Unfortunately, fewer than one-third of persons using cigarettes attempting to quit utilize evidence-based interventions like FDA-approved cessation medications or behavioral counseling.^3^ This shortfall in the use of interventions contributes to higher relapse rates, with successful quitters often needing up to 20 attempts in the absence of counseling and medications.^2^ Surveys show that from the patient’s perspective, physicians are advising patients to quit smoking routinely but not directing them to evidence-based treatment. Each year, the National Committee for Quality Assurance (NCQA) surveys members of every health plan to explore a variety of customer satisfaction measures.^4^ NCQA surveys for 2021 reveal that 44% of Commercial HMO members don’t receive counseling on how to quit, and fewer than 40% of patients have discussions about medications. These national survey data, repeated yearly, show that even when clinicians provide advice to quit, that intervention is often incomplete, lacking discussion of medication and cessation strategies. NCQA data may have significant response bias, which may lead to under-reporting of prevalence and overstatement of cessation interventions.^5^ Another perspective on tobacco cessation interventions comes from the National Health Interview Surveys, which stated that 63% of smokers reported physician advice to quit, and 6.2% were given a prescription in the most recent year sampled.^6^

Sadagna, et. Al. reported that in a large contemporary US registry, only 1 in 3 persons using cigarettes presenting for cardiology visits received smoking cessation assistance.^7^ Further, several reports looking at administrative data reveal significant variation and overall underperformance in the provision of smoking cessation services,^8,9,10^ which were attributed to barriers like time constraints, lack of training, and low reimbursement. In a Medicaid claims analysis across 37 states (where an average of 9.4% of people who smoke attempted to quit within the last year), cessation medication claims ranged from 0.2% to 32.9%, and an average of only 2.7% received cessation counseling (ranging from 0.1% to 5.6%).^10^ The majority of studies on tobacco cessation are focused specifically on smoking of cigarettes, but that literature can be applied to tobacco overall. The billing and coding of services for all forms of tobacco use are the same,**Error! Bookmark not defined**.^,21^ and the FDA-approved medications containing nicotine can be used interchangeably for any treatment of nicotine dependence, and non-nicotine medications appear to be effective as well.^11^

Advising a tobacco user to quit is a standard of care^12^ which is not usually captured in administrative data. Counseling persons using tobacco for three minutes or ten minutes is a billable service. Clinical practice guidelines support the delivery of a billable, evidence-based intervention at every clinical encounter, regardless of a patient’s readiness to change.^13^ Much of what is known about clinical performance in tobacco cessation is gleaned from survey data.^14^ Medical claims systems and EMR systems are likely to significantly under-report tobacco use status.^15,16^ Aside from the known challenges with electronic medical records-based claims and reimbursement,^17^ the potential revenue lost due to ineffective billing and underlying poor service delivery for cessation services remains largely unexplored. The lost revenue can be a marker for indicating sub-optimal interventions in health systems and quantifying the economic impact of missed tobacco cessation care.

This study aims to evaluate the potential revenue lost in tobacco cessation care due to inadequate billing practices. Inadequate billing reflects a mix of services not provided and services provided but not billed. By using lost revenue as a proxy, the study intends to measure the potential impact of applying systems-based approaches in population health for tobacco cessation. The objective is to develop an analysis framework for researching clinical data from multiple EMRs, increasing awareness in billing practices, and ultimately enhancing the effectiveness of tobacco cessation interventions.

## Methods

### Study Sample

In this cohort study, we used aggregated de-identified, patient-level health data to make population-level estimates of persons using tobacco across an 8-hospital health system, WellSpan Health. De-identified data from over 200 outpatient care locations across Pennsylvania and Maryland were mined to reveal the total number of primary care encounters eligible for tobacco cessation counseling. Over a three-year period, we examined patient’s tobacco use and whether tobacco cessation counseling was billed (CPT codes 99406 or 99407). Inclusion criteria are summarized in Table 1.

**Table 1.** Summary of Eligibility Parameters for Tobacco Cessation Care Billing.

| Criteria | Details<sup>a</sup> |
| --- | --- |
| Eligible Encounter Codes | Level 2-5 Evaluation and Management (E/M) codes, Qualifying Current Procedural Terminology (CPT) or Healthcare Common Procedure Coding System (HCPCS) codes |
| Tobacco Product Use | Use of any tobacco product, including but not limited to cigarettes, cigars, dissolvable, nicotine gels, hookah tobacco, pipe tobacco, roll-your-own tobacco, smokeless tobacco products, vapes, e-cigarettes, hookah pens, and other electronic nicotine delivery systems |
| Exclusion Criteria | Patients with limited life expectancy, including those receiving hospice services, and any patients with medical reasons that justify not screening for tobacco use or not providing tobacco cessation interventions.<sup>b</sup> |
| <sup>a</sup> List of CPT/HCPCS Codes \| CMS. Accessed February 6, 2024. https://www.cms.gov/medicare/regulations-guidance/physician-self-referral/list-cpt/hcpcs-codes |  |
| <sup>b</sup> Explore Measures & Activities. Accessed February 6, 2024. https://qpp.cms.gov/mips/explore-measures |  |
| Note: Tobacco cessation intervention can be performed by any healthcare provider, not necessarily the same provider who conducted the tobacco use screening.<sup>38</sup> |  |

Tobacco use status in electronic medical records will reflect the inconsistencies of the practices that collect this data.^16^ Despite best practices to screen for tobacco use at every visit and update the medical record accordingly,^18^ we recognized that many practices do not routinely capture tobacco use and may miss some tobacco users, and recent quitters might continue to be listed as current users.

### Measures

For data mining and inclusion criteria, we used custom data sessions within Epic’s SlicerDicer to model similar data capture and analysis across varying Electronic Medical Record (EMR) systems. Primary population filters in Epic’s SlicerDicer were used to obtain comprehensive patient-level and visit-level data. The timeframe was three years between January 1, 2021, and December 31, 2023. Persons using tobacco were identified using standard ICD-10 coding based on CMS standards.^**Error! Bookmark not defined**.21^ Adults aged 18 and over were identified through structured data elements in the EMR indicating tobacco use status (Table 1). Patients in hospice care were excluded as CMS does not pay for tobacco cessation for this population. This identification was derived from medical history records, active problem lists, or specific tobacco cessation billing codes. We used groupers to include sub-codes and decimal versions of ICD-10 codes for nicotine dependence (F17.*), tobacco use (Z72.0), and tobacco abuse (Z71.6); these details are in the appendix.

Regarding visit types, outpatient and telemedicine visits in the primary care service line were used exclusively, and data was reported in terms of unique patients and individual visits to provide context to the patient-to-visit ratio. Inpatient encounters, emergency department visits, and any visits by hospice patients were excluded.

To assess the prevalence of tobacco use in our patient cohort, was calculated by dividing the number of identified persons using tobacco by the total population. This calculated prevalence was then compared to the state health department’s tobacco use prevalence rates to ensure reliability.^19^

To determine and compute the billing rate, we examined the subset of the tobacco-using population billed for CPT codes 99406 or 99407 during the specified study period. Patients billed once or more (but less than eight) sessions per year proxied the frequency of cessation counseling. This data was segmented to differentiate between telemedicine (including audio-only) versus office encounters. To compare the revenue loss per patient and per visit, we calculated both billing rates by dividing the number of patients or visits billed for CPT codes 99406 or 99407 by the total number of identified tobacco users (i.e., patients) or their total number of encounters (i.e., visits).

To estimate the unrealized potential revenue loss, we identified persons using tobacco who were not billed for CPT codes 99406 or 99407. The reimbursement was calculated based on insurance type. Medicare reimburses up to eight cessation attempts within a 12-month period, with private plans and Medicaid typically adhering to these guidelines.^20^ Visits that were ineligible for reimbursement due to exceeding this limit or because of the patient’s hospice status were excluded from our analysis. The resulting number was then multiplied by the payer-specified reimbursement rate to estimate the financial impact of underbilling for tobacco cessation counseling services. The standard Medicare reimbursement rate for the three-minute counseling visit, 99406, was used for the calculation. The base rate of $14.97 was multiplied by 1.4 for commercial insurance and reduced by 70% for Medicaid.^21^ Since the majority of billable visits were for patients with Medicare, Medicaid, and commercial insurance types, these were reported. Other types of insurance and uninsured patients were removed from the estimates of missed opportunities.

Regarding demographics, data on five variables from patients 18 years and older who had an office visit or video visit at a primary care office between January 1, 2021, and December 31, 2023, was reported. The data for race and ethnicity were aggregated based on standards set by the U.S. Department of Health and Human Services and Office of Management and Budget.^22^ The sex of the patient was based on the legal sex documented in the patient’s chart. Age categories were based on the patient’s age at the time of the visit. Insurance data was based on the primary payer who covered the visit. Cumulative percentages of age group and insurance type exceeded 100% because patients may have had multiple encounters at different ages or with different insurance coverage during the study period. After building demographic criteria, we then grouped patients into segments based on whether they had an office visit or video visit. Cumulative percentages for visit type exceed 100% because patients may have visits using both modalities during the study period.

The WellSpan Health Institutional Review Board (IRB) waived full ethical review, categorizing the study as “non-human subjects research” (IRB: HE-2023-101) according to the regulations established by the DHHS and the FDA under 45 CFR 46.102 and 21 CFR 50.3, respectively.

## Results

A total of 584,631 unique patients who met the study inclusion criteria during the three-year sample period were identified. Demographics (Table 2) revealed a predominately white (83.4%) and non-Hispanic or Latino (88%) population. Over half (55.5%) were covered by commercial insurance, while Medicare accounted for 26.5% of patients and Medicaid 15.8%. There was a slightly higher representation of females (54.6%). Patients were distributed among four age groups, with the majority being older adults (22.1% aged 18-29 years, 26.2% aged 30-44 years, 34.4% aged 45-64 years, and 26.9% aged 65 years and above). The primary visit modality was in-office (91.1% of patients had at least one office visit), while 13.6% of patients had at least one video visit (4.6% had both).

**Table 2.** Patient Demographics by Visit Type, WellSpan Health (January 1, 2021 to December 31, 2023)

|  | Office | % | Telemedicine | % | Total Patients <sup>a</sup> | % |
| --- | --- | --- | --- | --- | --- | --- |
| **Race** |  |  |  |  |  |  |
| White | 444,142 | 83.4% | 67,570 | 85.3% | 487,617 | 83.4% |
| Other | 36,957 | 6.9% | 5,419 | 6.8% | 40,005 | 6.8% |
| Black or African American | 25,750 | 4.8% | 3,920 | 4.9% | 27,724 | 4.7% |
| Unknown, declined, or not reported | 18,301 | 3.4% | 1,394 | 1.8% | 21,115 | 3.6% |
| Asian | 6,276 | 1.2% | 761 | 1.0% | 7,059 | 1.2% |
| American Indian, Alaska Native, Native Hawaiian and Other Pacific Islander | 1,018 | 0.2% | 169 | 0.2% | 1,111 | 0.2% |
| **Ethnicity** |  |  |  |  |  |  |
| Not Hispanic or Latino | 469,761 | 88.2% | 71,716 | 90.5% | 514,657 | 88.0% |
| Hispanic or Latino | 38,485 | 7.2% | 5,659 | 7.1% | 41,633 | 7.1% |
| Unknown, declined, or not reported | 24,198 | 4.5% | 1,858 | 2.3% | 28,341 | 4.8% |
| **Legal sex <sup>b</sup>** |  |  |  |  |  |  |
| Female | 292,413 | 54.9% | 53,592 | 67.6% | 319,039 | 54.6% |
| Male | 239,943 | 45.1% | 25,619 | 32.3% | 265,498 | 45.4% |
| **Age <sup>c</sup>** |  |  |  |  |  |  |
| 18-29y | 119,222 | 22.4% | 18,305 | 23.1% | 129,076 | 22.1% |
| 30-44y | 141,613 | 26.6% | 29,582 | 37.3% | 153,093 | 26.2% |
| 45-64y | 182,827 | 34.3% | 29,394 | 37.1% | 201,231 | 34.4% |
| 65+y | 140,836 | 26.5% | 13,097 | 16.5% | 157,133 | 26.9% |
| **Insurance type <sup>d</sup>** |  |  |  |  |  |  |
| Commercial (Highmark, Blue Cross, etc.) | 294,719 | 55.4% | 48,078 | 60.7% | 324,457 | 55.5% |
| Medicare | 140,399 | 26.4% | 13,900 | 17.5% | 155,162 | 26.5% |
| Medicaid | 87,684 | 16.5% | 16,457 | 20.8% | 92,285 | 15.8% |
| Government/Tricare | 9,189 | 1.7% | 1,035 | 1.3% | 10,042 | 1.7% |
| Other | 64,605 | 12.1% | 3,488 | 4.4% | 125,421 | 21.5% |
| **Total** | **532,444** | **91.1%** | **79,233** | **13.6%** | **584,631** | **100.0%** |

<sup>a</sup> Cumulative percentages for visit types exceed 100% as patients may have both an office visit and video visit during the study period.
<sup>b</sup> Percentages are calculated based on tallies of patients' legal sex since these values remain consistent over time
<sup>c</sup> Age values can show discrepancies of approximately 5% between cohorts, primarily because some ages are recorded in months and may not be accurately accounted for in the EMR. Additionally, certain ages can change during the measurement time frame, so cumulative percentages exceed 100%.
<sup>d</sup> SlicerDicer can only measure proportions of encounters associated with the financial payer class, making them unavailable for regression analysis. Separating self-pay data from cost-sharing (co-pays and deductibles) can be challenging, resulting in imprecise measurements, as patients can switch payers within the study period. Notably, this challenge can be exacerbated in the blended group, where patients receive both types of care. In the case of Medicare and Medicaid, there might be overlap among patients with both payer types. The key insight from the payer data is the comparable distribution of percentages across exposure groups.

A total of 75,115 persons using tobacco were identified over the study timeframe, or 12.8% of the patient population (Table 3). This was comparable to the state-specific figures on tobacco use recorded at 14.4% for Maryland and 18.5% for Pennsylvania.^23^ These patients had 507,656 total primary care encounters over the timeframe potentially eligible for receiving tobacco cessation counseling. We found only 1,277 patients, or 1.7% of eligible patients, were billed for tobacco cessation counseling during the study period.

**Table 3.** Primary Care Tobacco Cessation Counseling by Year.

| Category | 2021 (%) | 2022 (%) | 2023 (%) | Total (%) | Average (%) |
| --- | --- | --- | --- | --- | --- |
| **Patients** |  |  |  |  |  |
| Total | 405,485 | 380,204 | 375,704 | 584,631 | 387,131 |
| Persons using tobacco | 43,695<br>(10.8%) | 44,285<br>(11.6%) | 42,385<br>(11.3%) | 75,115<br>(12.8%) | 43,455<br>(11.2%) |
| Persons using tobacco billed for cessation | 622 (1.4%) | 545 (1.2%) | 218 (0.5%) | 1,277 (1.7%) | 462 (1.1%) |
| **Visits** |  |  |  |  |  |
| Persons using tobacco, total primary care encounters | 169,215 | 172,688 | 165,753 | 507,656 | 169,219 |
| Persons using tobacco, encounters billed for cessation | 731 (0.4%) | 596 (0.3%) | 230 (0.1%) | 1,557 (0.3%) | 519 (0.3%) |
| Encounters with other insurance type or no insurance | 11,889 | 11,067 | 9,631 | 32,587 | 10,862 |
| Non-reimbursable encounters* | 24,047 | 24,508 | 22,459 | 71,014 | 23,671 |
| Unbilled encounters | **132,548** | **136,517** | **133,433** | **402,498** | **134,166** |
| * This number reflects visits that exceed the maximum reimbursable visits for our 3 payers of interest. For example, Medicare covers up to 8 cessation attempts per year, so Medicare patients with more than 8 visits in a year will only count as having 8 eligible visits. |  |  |  |  |  |

In terms of encounters, tobacco users had 507,656 office visits (Table 2), and only 1,557 (0.3%) were billed for cessation services. Over the 3-year timeframe, patients with Medicare, Medicaid, and commercial insurance had a total of 402,498 potentially eligible visits that were not billed for tobacco cessation counseling (Table 4). Of the eligible opportunities to provide and bill for tobacco cessation services, 97% appear to have been missed. The estimated potential reimbursement for tobacco cessation care, if provided and billed consistently, was $5,947,018.13 over three years (Table 4).

**Table 4:** Estimated Reimbursement Based on Payer Type.

| Payer | 2021 Eligible Visits<br>(est. reimbursement) | 2022 Eligible Visits<br>(est. reimbursement) | 2023 Eligible Visits<br>(est. reimbursement) | Total Eligible Visits<br>(est. reimbursement) | Avg. Eligible Visits<br>(est. reimbursement) |
| --- | --- | --- | --- | --- | --- |
| Commercial<br>\$20.96 | 35,864<br>(\$751,637.71) | 36,359<br>(\$762,011.92) | 35,471<br>(\$743,401.22) | 107,694<br>(\$2,257,050.85) | 35,898<br>(\$752,350.28) |
| Medicare<br>\$14.97 | 41,986<br>(\$628,530.42) | 44,995<br>(\$673,575.15) | 46,779<br>(\$700,281.63) | 133,760<br>(\$2,002,387.20) | 44,587<br>(\$667,462.40) |
| Medicaid<br>\$10.48 | 54,698<br>(\$573,180.34) | 55,163<br>(\$578,053.08) | 51,183<br>(\$536,346.66) | 161,044<br>(\$1,687,580.08) | 53,681<br>(\$562,526.69) |
| **Total** | **132,548**<br>**(\$1,953,348.47)** | **136,517**<br>**(\$2,013,640.15)** | **133,433**<br>**(\$1,980,029.51)** | **402,498**<br>**(\$5,947,018.13)** | **134,166**<br>**(\$1,982,339.38)** |

## Discussion

This study found a significant gap between persons using tobacco and billed encounters for tobacco cessation care. Leveraging a large health system’s aggregated patient data, the unrealized revenue highlights two concerns in tobacco cessation care: underbilling (when billable care may have been rendered), and failure to provide care to eligible patients. The findings highlight the importance of improving billing practices, even in telemedicine, which has become increasingly common and can represent a significant proportion of the population (13.6% of our study’s participants, as shown in Table 2). Given the prevalence of tobacco use, the impact of missed opportunities to help patients quit the use of tobacco is profound.

Our reported billing rate—1.7 % overall, is consistent with other studies.^10^ In a recent study of an EMR-enabled smoking treatment program, the pre-intervention baseline was 2.1%.^24^ But survey data supports persons using cigarettes’ affirmation that their doctors are providing tobacco cessation services at a much higher rate.^25,14^ This highlights the importance of improving billing practices, even in telemedicine, which has become increasingly common and can represent a significant proportion of the population (13.6% of our study’s participants, as shown in Table 2). The billing rates for relevant CPT codes (99406 or 99407) were considerably low, despite the significant portion of the eligible population that could benefit from cessation interventions,^26^ as indicated by the overall visit-level billing rate of only 0.3% (Table 3).

It is important to note in our analysis that eligibility for tobacco cessation services was not static. This reflects the reality that patients’ smoking status changes over time, impacting their potential eligibility for cessation services within the study timeframe. Methodologically, if the EHR indicated the patient had quit, they would no longer trigger eligibility for cessation interventions in subsequent visits. However, our use of aggregate data does not allow for distinguishing between patients moving in and out of the health system from patients who drop off the eligibility threshold. Nonetheless, the billing rate remained consistently low at around 0.3% (Table 3) despite patient and visit numbers changing over the study timeframe. Additionally, by excluding uninsured and other unmeasured insurance from our calculations, over the three years, the potential revenue lost is still estimated at $5,947,018.13, demonstrating a substantial financial impact. This translates to an average $1,982,339.38 potential revenue loss per year. Moreover, this is potential revenue that is not reinvested into frontline clinicians and their patients— blunting the impact of primary prevention.

Many studies explore the barriers to systemic implementation of tobacco cessation in detail,^27,28^ however, there is a notable gap in the literature concerning the reimbursement perspective. The most frequently reported perceived barriers to providing cessation services were time constraints, high workload, lack of training, lack of reimbursement of smoking cessation interventions, and the physician’s own smoking status. ^**Error! Bookmark not defined**.^ Low outcome expectancy, lack of self-efficacy, fear of losing patients, being uncomfortable discussing smoking hazards, and dissatisfaction with professional activity were also reported.^**Error! Bookmark not defined**.,29^ Other studies have produced similar results in assessing effective smoking cessation intervention in primary care^30^ and tobacco treatment guideline use and predictors among U.S. physicians by specialty.^31^

Our study’s results call for a strategic healthcare policy review by every system, tracking the prevention and screening opportunity from encounter to outcome and reimbursement. In seeking to optimize systems performance in identifying persons using tobacco, accurate documentation of a visit, shaping additional treatment and follow-up, and capturing appropriate billing, AI-driven clinical billing systems have a clear potential role.^32^ Although there is a cost to systems change, the return on investment from tobacco cessation is well established.^33^ Additionally, the study highlights the necessity of raising awareness among medical providers and their leadership about adopting systems-based solutions to tobacco cessation with attention to documentation and billing. Well-established paradigms for systems change have been developed and evaluated.^34^

These include elements of EMR and claims system design, training, and program evaluation.^35,36,37^ Through these actions, it is reasonable to presume improvement in tobacco cessation interventions, increased revenue for medical practices and reduced burden of tobacco-related diseases.

Our study provides valuable insight to providers measuring the potential impact of applying value-based approaches in population health for tobacco cessation. Ultimately, this research emphasizes the critical need for systemic changes that not only foster financial efficiency but also prioritize and effectively deliver public health interventions. Implementing such changes could lead to a paradigm shift in healthcare delivery, aligning financial incentives with public health goals and elevating the quality of patient care.

### Limitations

We acknowledge several limitations of our study. First, while we provide a general framework for providers to measure the impact of undelivered tobacco cessation, our results have limited generalizability to other parts of the country outside the northeast United States as they are from one health system. A second limitation is that the estimated revenue loss is conservative.

Tobacco use is not adequately captured in EHRs, and the reimbursement estimate was based on only the three-minute counseling visit (CPT 99406). Given the national prevalence of tobacco use is 18.7%,^1^ the reported results of missed opportunities are likely understated. Third, we only considered primary care encounters. This means we excluded any other outpatient specialties that might also be offering tobacco cessation counseling. Because we filtered our data by primary care, there may be a greater burden of unrealized potential that we did not measure. The current report is based entirely on EMR indicators of tobacco use and claims data and does not capture the full range of tobacco cessation services such as prescriptions and/or referral to quit lines. Future studies should attempt to capture this type of information to better distinguish between underbilling and opportunities completely missed. Finally, our study is limited by the wide variation in payer reimbursement, even for the same CPT codes, along with the cautious estimates we employed when assessing the three payers. Actual reimbursement varies by region and contract, which should be considered when interpreting our findings. Some contracts with insurers may be based on capitation, and the services may not be specifically reimbursable. However, the return on investment from smoking cessation is well established, and the rate of reimbursement underestimates the future avoided medical expense.

## Conclusion

This study provides evidence of the untapped potential revenue for consistent and systematic tobacco cessation care. The marked discrepancy between eligible and billed cessation services points to either a possible broader trend of underutilizing public health interventions, or a lack of adequate validation in tobacco cessation care delivery. This highlights an opportunity to apply systems-based approaches to tobacco cessation care to improve patient outcomes and economic returns.

## Data Availability

All data produced in the present study are available upon reasonable request to the authors

## Notes

All authors have contributed significantly to the work, meet ICMJE criteria, and approve the manuscript for submission. There are no conflicts of interest to disclose, financial or otherwise, and all study procedures were conducted under ethical standards. No funding was received for this study.

### Competing Interest Statement

The authors have declared no competing interest.

### Funding Statement

This study did not receive any funding

### Author Declarations

The WellSpan Health Institutional Review Board (IRB) waived full ethical review, categorizing the study as "non-human subjects research" (IRB: HE-2023-101) according to the regulations established by the Department of Health and Human Services (DHHS) and the Food and Drug Administration (FDA) under 45 CFR 46.102 and 21 CFR 50.3, respectively

### Summary of Updates

Summary of the major changes made: 1.Clarification of Billing Rates: We have revised the abstract and methods section to clearly define and calculate both per-encounter and per-patient billing rates, addressing the confusion highlighted by peer reviewers. 2.Methods Explanation: Enhanced the description of our methodology to better explain how patients were identified as smokers, including the dynamic identification process and the handling of eligibility throughout the study period. 3.Revenue Calculations: Improved the explanation of our revenue loss calculations, ensuring a clearer distinction between services provided but not billed and those not provided. 4.Limitations: Added more detailed discussions on systemic barriers to effective tobacco cessation interventions and acknowledged the limitations of our data, including the exclusion of certain outpatient specialties and potential underreporting due to EMR inconsistencies. 5.Updated References and Data: Incorporated recent studies and data to support our findings and revised several sections for improved readability and accuracy.

